# Epidemic characteristics of respiratory viruses in hospitals in a Chinese city during the SARS-CoV-2 epidemic

**DOI:** 10.1101/2020.10.06.20207589

**Authors:** Weihua Yang, Jian Chen, Mingjie Xu, Jun Wang, Huanjie Li, Yunshan Wang

## Abstract

The SARS-CoV-2 virus first broke out in China in early 2020. The early symptoms of COVID-19 are similar to those of influenza. Therefore, during the epidemic, patients with similar symptoms will be tested for multiple pathogens at the same time. In order to control the spread of SARS-CoV-2, China has taken many measures. Under this special situation, have the types and epidemic characteristics of respiratory viruses changed? The nucleic acid test results of influenza A virus, influenza B virus and respiratory syncytial virus, as well as the antibody test results of 8 common respiratory viruses of Jinan Central Hospital were collected before and after the occurrence of SARS-CoV-2, and age distribution and time distribution characteristics were statisticed. Furthermore the epidemiological characteristics of this new virus before and after the SARS-CoV-2 epidemic was compared. In the early stage of the SARS-CoV-2 epidemic, influenza A, influenza B and respiratory syncytial virus nucleic acid test samples were large, and the positive rate of the three viruses was high. After that, the sample size and positive rate decreased significantly. No co-infection of SARS-CoV-2 and other viruses was found in our hospital. The sample size before the SARS-CoV-2 outbreak was larger than that after the outbreak, but the positive rate of the outbreak was lower than that after the outbreak. And the infection rate of children decreased in the middle and late stages of the epidemic. This is because since January 23, in order to prevent the spread of the new crown epidemic, my country has adopted measures such as wearing masks, not gathering together, and quarantining at home. This not only prevents the spread of the new crown virus, but also prevents the common respiratory tract. The spread of the virus has reduced the incidence of residents.

---

Acute respiratory infections are an important public health problem worldwide, causing predictable morbidity and mortality in all age groups^1^. The average infection frequency of children is 2 to 3 times that of adults^2^, and there are more than 200 types. Respiratory viruses can cause acute respiratory infections. Human respiratory syncytial virus, adenovirus, parainfluenza virus, influenza A virus, influenza B virus, chlamydia, and mycoplasma are the most common pathogenic microorganisms of acute respiratory infections in children, accounting for about 70% of acute respiratory infections^3, 4^. The **SARS-CoV-2** first broke out in China in early 2020^5^. In order to control the spread of the virus, China adopted measures such as wearing masks, suspension of work and school, home isolation, and strengthening of virus testing. Under this special situation, have the types and epidemic characteristics of respiratory viruses changed? Therefore, the results of the respiratory virus test in Jinan Central Hospital before and after the occurrence of the new coronavirus were compared.

## 1. Materials and Methods

### 1.1 Samples

This study included samples from December 23, 2019 to March 6, 2020. Patients with acute respiratory infections in children and adults who are clinically expected to have the following 2 symptoms: fever, sore throat, cough, sputum, shortness of breath, Abnormal lung auscultation, shortness of breath, chest pain. Take a nasopharyngeal swab or venous blood from the patient.

### 1.2 Detection method

#### 1.2.1 Nucleic acid detection method

363 cases of nasopharyngeal swabs from January 25 to March 6 were collected for nucleic acid detection. Extract viral RNA and detect whether it contains influenza A virus, influenza B virus and respiratory syncytial virus by real-time quantitative PCR. Nucleic acid tests for **SARS-CoV-2** are performed on positive samples, and the three respiratory viruses are tested on positive samples for the new coronavirus to determine whether **SARS-CoV-2** and these three viruses are simultaneously infected.

#### 1.2.2 Antibody detection method

1449 cases of venous blood were collected from December 23 to February 24 for antibody detection, 808 cases were from December 23 to January 22, and 641 cases were fromJanuary 23 to February 24. After centrifugation of venous blood, immunofluorescence was used to detect antibodies to 8 common respiratory viruses (Parainfluenza virus, Mycoplasma pneumoniae, Legionella, Influenza A, Influenza B, Chlamydia pneumonia, Adenovirus, Respiratory syncytial virus) in serum.

### 1.3 Statistical method

Count data is expressed in [n(%)], using χ2 test; P<0.05 indicates that the difference is statistically significant.

## 2. Result

### 2.1 Nucleic acid testing of SARS-CoV-2 suspected patients

#### 2.1.1 Overall positive rate

A total of 363 samples were tested and 59 were positive, with a positive rate of 16.25%. Among them, FluB had the highest positive rate, 7.99% (29/363); followed by FluA, with a positive rate of 5.23% (19/363); respiratory syncytial virus RSV had the lowest positive rate, with 2.20% (8/363). These 363 samples were tested negative for **SARS-CoV-2**nucleic acid. During this period, our hospital detected a total of 5 positive samples of **SARS-CoV-2** nucleic acid, which were all negative after FluA, FluB and RSV nucleic acid tests.

#### 2.1.2 Time distribution

Figure 1 and Figure 2 showed,from January 25 to February 4, the sample size is large, and the positive rate of the three viruses is high. After that, the sample size and positive rate were significantly reduced. From February 6 to February 20, the number of consultations was low, and no positive was detected. After February 21, the number of doctors increased, but the positive rate of the three viruses was low. These 363 samples were tested negative for **SARS-CoV-2** nucleic acid. During this period, our hospital detected a total of 5 positive samples of **SARS-CoV-2** nucleic acid, which were all negative after FluA, FluB and RSV nucleic acid tests.

**Figure 1.**
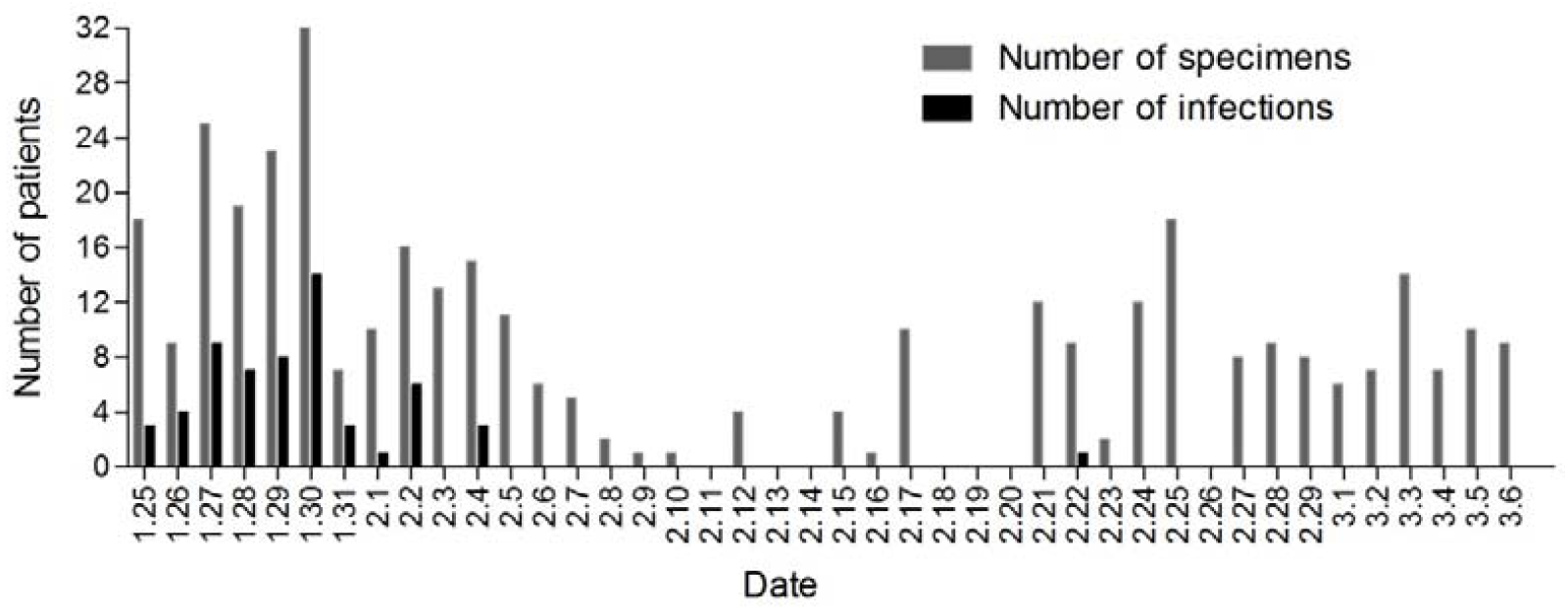
Number of positive samples and total number of samples for nucleic acid testing

**Figure 2.**
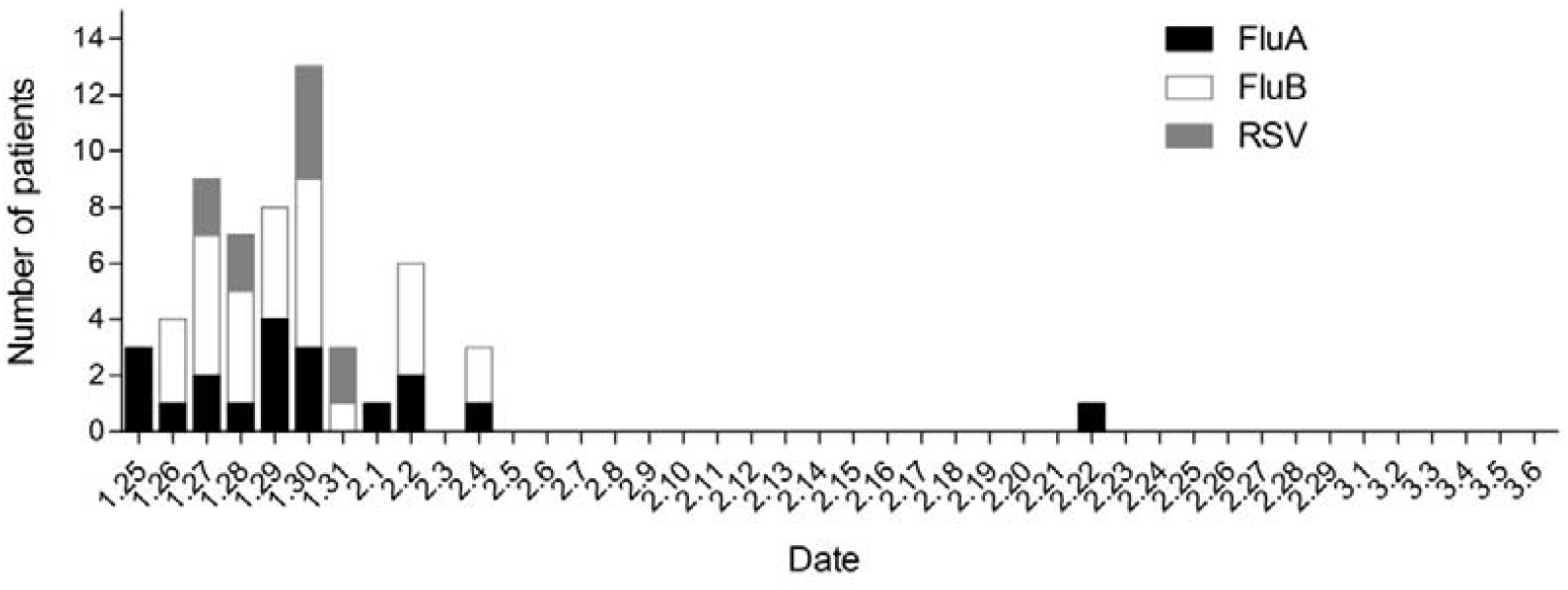
Daily distribution of three virus-positive specimens (January 25-February 4)

### 2.2 Immunofluorescence method to detect 8 common respiratory pathogens

Taking January 23 as the demarcation line of the epidemic, 808 samples were tested one month before the outbreak, and positive samples accounted for 75.2% (608/808), and the combined infection rate of the two viruses was 15.6%; one month after the outbreak, 641 samples were tested positive The samples accounted for 94.8% (607/641), and the combined infection rate of the two viruses was 9.6%. Among the 8 pathogens, the top three were MP, LGB and LDB. The positive rates of MP were 57.8% (475/808) and 61.2% (392/641) (P>0.05), and the positive rates of LGB were 23.6% (191/808) and 35.7% (229/641) (P>0.05), the positive rates of LDB were 5.3% (43/808) and 4.4% (28/641), (P<0.05), positive for several other pathogens The rates are all less than 10%, as shown in Figure 3.

**Figure 3.**
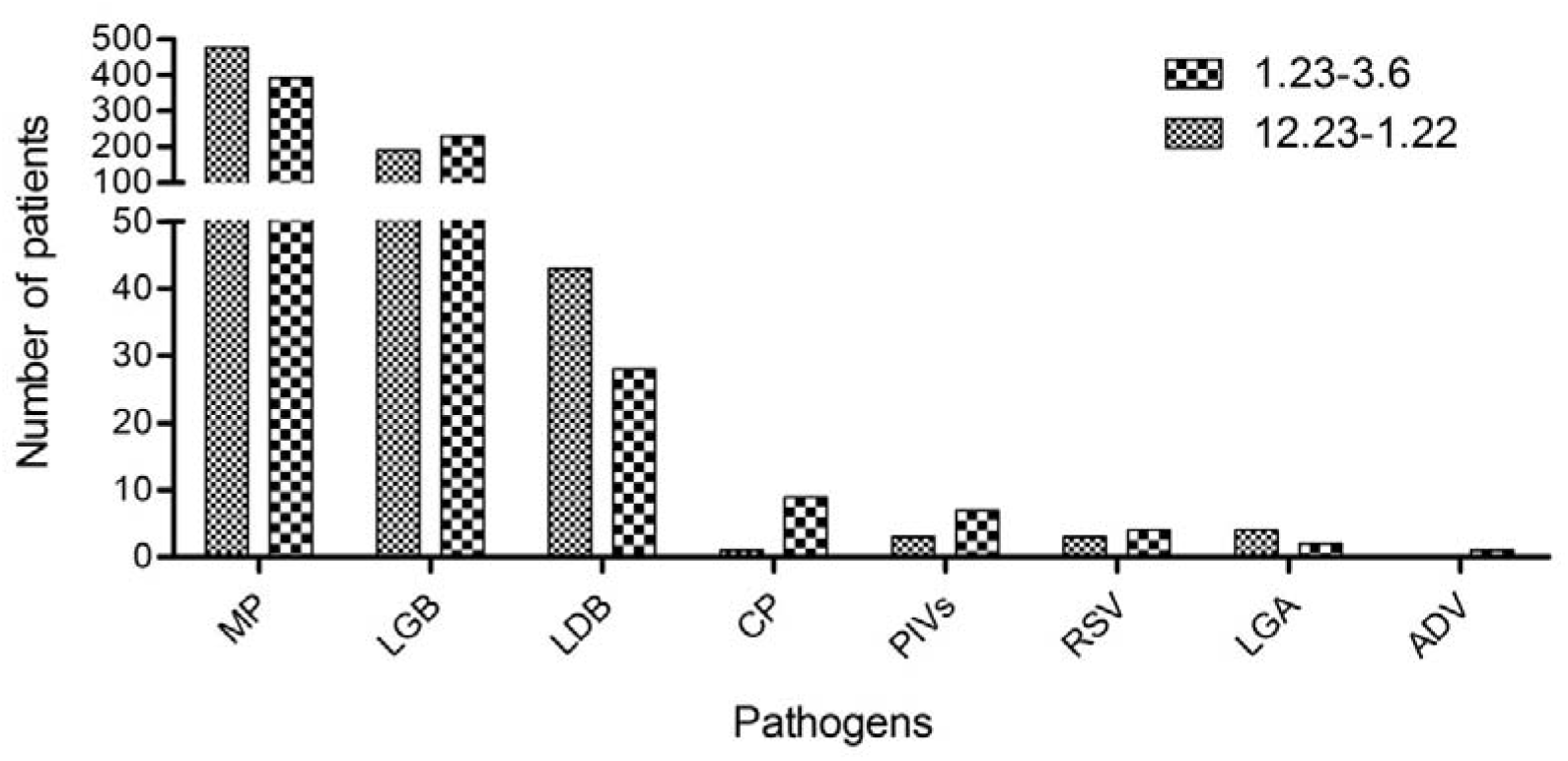
The occurrence of 8 respiratory pathogens before and after the new crown epidemic

### 2.3 Distribution of 8 viruses in different age groups

The positive rate of MP in the children’s age group and the 21-30 age group is over 50%, accounting for 57.4% and 58.6% of the pathogens in this group respectively; the positive rate of LGB in the 31-40, 61-70 and older age groups More than 50%, respectively, accounted for 55.6%, 54% and 52.8% of the pathogens in this group; LDB is most likely to occur in people over 80 years old, with a positive rate of 9.7%. See Figure 4.

**Figure 4.**
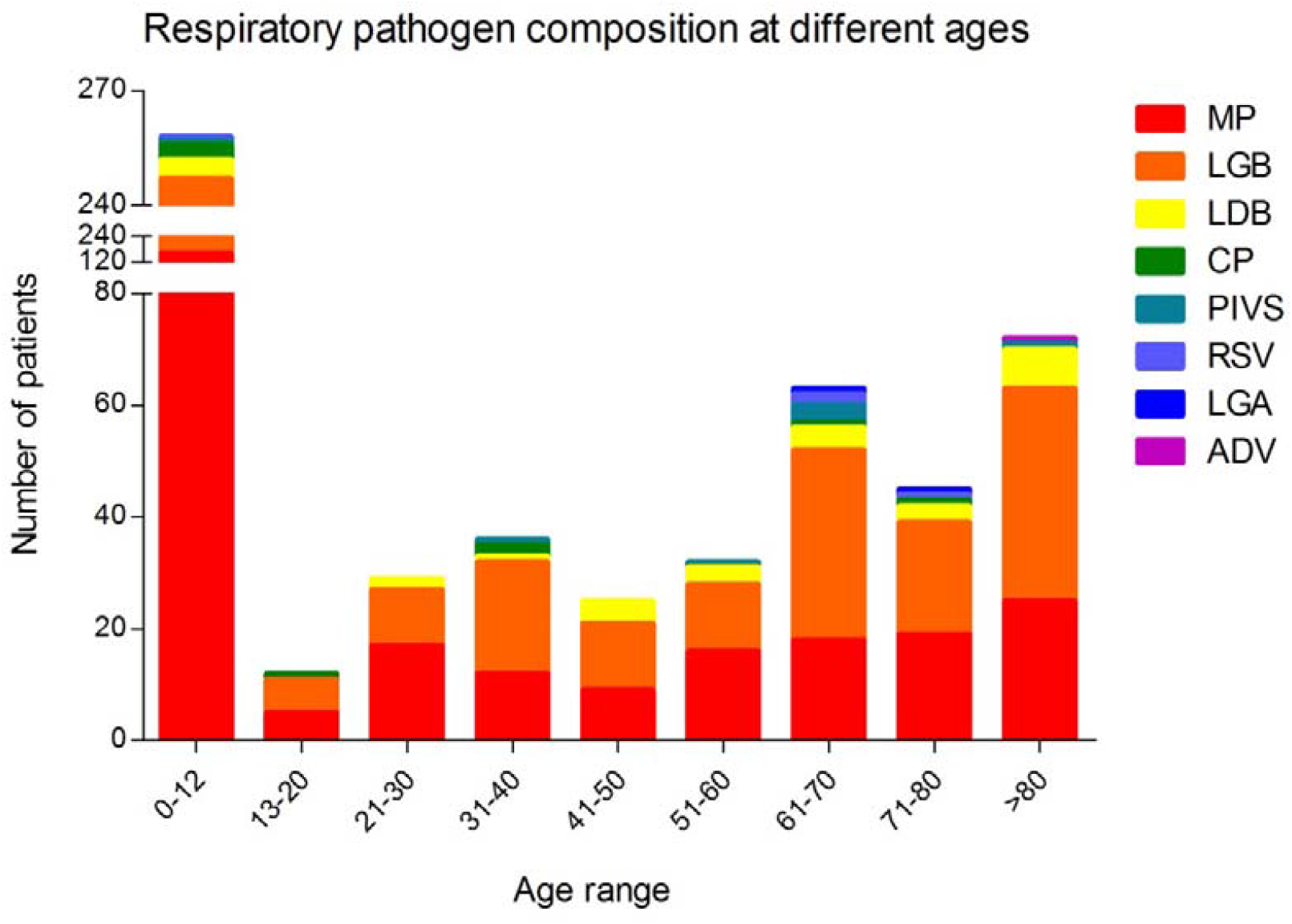
Distribution of 8 viruses in different age groups

### 2.4 Epidemic characteristics of influenza B virus

Figure 5 showed before the outbreak (before January 23) and at the beginning of the outbreak (January 23-February 3), the incidence of influenza B in children was very high, accounting for more than 50% of the total number of positives almost every day. As time progresses, After February 5, the child infection rate decreased significantly, and until February 23, except for February 18, it was below 40% every day.

**Figure 5.**
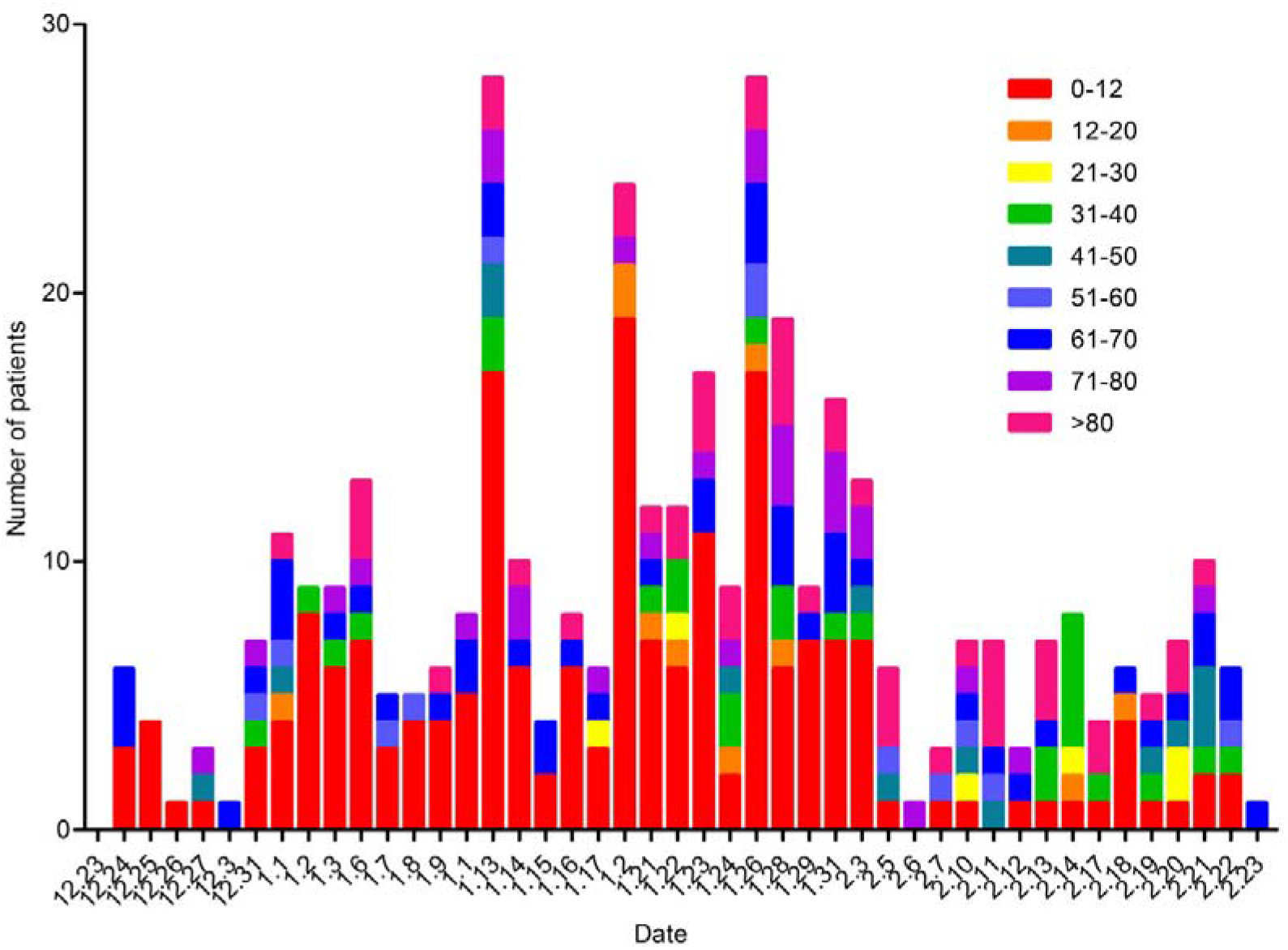
The distribution of influenza B viruses at different ages and times

## 3. Discussion

The typical clinical manifestations of COVID-19 are mainly fever, dry cough, and fatigue^6^. Early symptoms include fever, cough, throat swelling and pain, and some patients have upper respiratory tract catarrhal symptoms including nasal congestion and runny nose^7^. Easily confused with influenza. In the early stage of the epidemic, in order to rule out COVID-19, our hospital first tested the nucleic acid of influenza A, influenza B and respiratory syncytial virus, and focused on the detection of SARS-CoV-2 nucleic acid for patients with negative nucleic acids of these three viruses. At the beginning of the SARS-CoV-2 epidemic, the nucleic acid test samples were large, and the positive rate of the three viruses was high. After that, the sample size and positive rate were significantly reduced. As the epidemic spreads, all suspected patients will be tested for nucleic acid of the new coronavirus, resulting in a decrease in the amount of the nucleic acid testing for influenza A, influenza B and respiratory syncytial virus. With seasonality, the influenza peak is over, and the positive rate of these three viruses was reduced. A total of 5 samples with positive nucleic acid of SARS-CoV-2 were detected in our hospital, all of which were FluA, FluB and RSV nucleic acid negative. Furthermore,all influenza A, influenza B and respiratory syncytial virus positive samples are SARS-CoV-2 negative. No co-infection of SARS-CoV-2 and other viruses has been found in our hospital.

Acute respiratory infection is a serious health problem with a high incidence. Children are susceptible and high-incidence groups of this disease. We have counted the antibody test data of 8 kinds of respiratory viruses, showing that the sample size before SARS-CoV-2 outbreak was larger than that after the outbreak, but the positive rate of the outbreak was lower than after the outbreak. Although the main pathogens have not changed, the top three are MP, LGB and LDB, but the distribution of each age group has changed. For example, before and at the beginning of the outbreak, the incidence of influenza B in children is very high. Over time, the infection rate of children is significantly reduced. This is because since January 23, in order to prevent the spread of the new crown epidemic, my country has adopted measures such as wearing masks, not gathering, and quarantining at home, which not only blocked the spread of the new crown virus, but also blocked the spread of common respiratory viruses, reducing the incidence of residents.

## Data Availability

All data referred to in the manuscript and note links below is availability.

## References

1. Chun Lin HC, Ping He,Yazhen Li,Changwen Ke,Xiaoyang Jiao. Etiology and characteristics of community-acquired pneumonia in an influenza epidemic period. Comparative Immunology, Microbiology and Infectious Diseases. 2019;64:153–158.

2. Williams BG GE, Boschi-Pinto C. Estimates of world-wide distribution of child deaths from acute respiratory infections. Lancet Infect Dis. 2002(2):25–32.

3. Brittain-Long R NS, Olofsson S. Multiplex real-time PCR for detection of respiratory tract infections. J Clin Virol. 2008(41):53–56.

4. Kusel MM DKN, Holt PG. Role of respiratory viruses in acute upper and lower respiratory tract illness in the first year of life:a birth cohort study Pediatr Infect Dis J. 2006(25):680–686.

5. Qun Li MM, Xuhua Guan, Peng Wu, et al. Early Transmission Dynamics in Wuhan, China, of Novel Coronavirus–Infected Pneumonia. The New England Journal of Medicine. 2020(382):1199–1207.

6. Cheng Lee T-IH, Yi-Hsiang Huang. The typical clinical manifestations of COVID-19. J Chin Med Assoc. 2020;83(6):521–523.

7. Ling Mao HJ, Mengdie Wang, Yu Hu, Shengcai Chen, et al. Neurologic Manifestations of Hospitalized Patients With Coronavirus Disease 2019 in Wuhan, China JAMA Neurol. 2020;77(6):683–690.

